# Late start of eating is linked to hyperinsulinemia, insulin resistance, and increased body fat in prediabetes

**DOI:** 10.64898/2026.01.01.25342809

**Authors:** Beeke Peters, Noelle Fröhlich, Jürgen Machann, Ulrike Dambeck, Caroline Honsek, Christiana Gerbracht, Anna Sachno, Margrit Kemper, Stefan Kabisch, Andreas Fritsche, Knut Mai, Andreas F.H. Pfeiffer, Olga Pivovarova-Ramich

## Abstract

**Background:** Prediabetes is associated with an increased risk of progression to type 2 diabetes. While dietary interventions in prediabetes traditionally focus primarily on energy and macronutrient intake, the role of eating timing has recently been highlighted. This study aimed to examine the relationships between eating timing pattern and glycaemic parameters in prediabetes.

**Methods:** 297 individuals with prediabetes, i.e. impaired fasting glucose or impaired glucose tolerance (age 59.5 y, BMI 31.3 kg/m^2^), underwent detailed metabolic phenotyping including an oral glucose tolerance test and magnetic resonance imaging/spectroscopy. Parameters of temporal eating pattern (eating timing, daily calorie distribution, and meal number) were extracted from four-day food records. Eating start (ES) was defined as the start of the first caloric event of the day.

**Results:** Among eating timing parameters, ES showed the most associations with glycaemic traits. Each later hour of ES was associated with a 1.08 mU/l higher fasting insulin (95% CI, 0.40 to 1.77 mU/l), a 16.86 mU/l higher 120-min insulin (95% CI, 5.62 to 28.10 mU/l), a 1147 mU/l*min higher AUC insulin (95% CI, 390 to 1904 mU/l*min), a 0.32 units higher HOMA-IR (95% CI, 0.14 to 0.50), and a −0.18 units lower Matsuda insulin sensitivity index (95% CI, −0.32 to −0.04) after adjustment for age, sex, and daily energy intake. Glycemic associations remained after additional adjustment for food composition as well as body fat percentage, which was increased in individuals with later ES (0.81% per ES hour; 95% CI, 0.27 to 1.34%). No associations of eating timing with visceral or liver fat were found.

**Conclusion:** Late ES is associated with estimates of hyperinsulinemia and lower insulin sensitivity in prediabetes, independent of energy intake, food composition, and body fat. Earlier ES might relate to improved glucose metabolism and lower the risk of progression to diabetes.

## 1. Introduction

The pandemic of diabetes mellitus affects millions of people worldwide. Type 2 diabetes (T2D), the most common form, is preceded by prediabetes, characterized by increasing blood glucose levels which does not yet achieve the diagnostic criteria for diabetes (1). Dietary interventions, aiming to impede the progression of prediabetes to diabetes, have traditionally focused on energy and macronutrient intake. Recent research has highlighted the importance of eating timing in managing of blood glucose levels and potentially in the prevention of T2D (2; 3). This fact is based on the diurnal variation of metabolic functions resulting in different metabolic responses to the same food consumed at different times of the day (4; 5).

Eating timing has emerged as a significant factor in glucose metabolism and insulin sensitivity (2; 6). Cross-sectional studies have shown that early eating patterns aligning with individual circadian rhythms are associated with lower body weight and metabolic health (3; 6-8). In interventional studies, eating earlier in the biological day resulted in greater reductions in body weight, BMI, and waist circumference, and improved glycaemic control. For instance, early time-restricted eating (TRE), limiting food intake to a 6-10 hour window early in the day, improved glucose tolerance and insulin sensitivity in several studies (9–12), although other research did not confirm this finding (13; 14) and numerous cofounders (longer fasting, fewer meals, lower calorie intake, and possibly different meal composition) has to be considered. In contrast, late and night eating showed an association with increased risk of obesity, cardiovascular diseases, and related traits such as higher BMI and body fat (6; 15-19).

Timing of specific eating events, particularly breakfast, appears to play a crucial role in glycaemic control. Most previous studies suggest that skipping breakfast has been linked to a higher risk of developing T2D as partly mediated by BMI (20), while the earlier breakfast timing was related to the lower T2D incidence (21). Nevertheless, metabolic risks of dinner skipping were also described (22). This suggests that not only timing of meals but also daily pattern of calorie distribution may support managing blood glucose levels in diabetes (23; 24). Additionally, the number of eating events and the duration of overnight fasting have been investigated for their potential impact on glucose regulation and diabetes risk (3; 25; 26). Nevertheless, it remains insufficiently studied how temporal eating pattern and its specific parameters, such as timing, frequency, and diurnal calorie distribution, contribute to the parameters of glucose metabolism in prediabetes and generally to diabetes risk. Therefore, this study aimed to examine relationships between eating timing and parameters of glucose metabolism in individuals with prediabetes.

## 2. Methods

### 2.1. Study participants and study protocol

This cross-sectional analysis included baseline data of 297 individuals with prediabetes obtained in two clinical trials, the Prediabetes Lifestyle Intervention Study (PLIS, NCT01947595, n=174) and the Diabetes Nutrition Algorithm - Prediabetes (DiNA-P, NCT02609243, n=123). Complete details of these trials have been described elsewhere (27–29). Both trials were conducted in accordance with the principles of the Declaration of Helsinki and Good Clinical Practice guidelines. The study protocol was approved by the ethics committee of the Charité-Universitätsmedizin Berlin. All patients provided written informed consent before enrollment.

Data included in this study were collected at the German Institute of Human Nutrition Potsdam-Rehbruecke and at the Charité-Universitätsmedizin Berlin between 2013 and 2019. Inclusion criteria in both trials were age between 18 and 75 years, BMI < 45 kg/m², and diagnosis of impaired fasting glucose (IFG; fasting blood glucose 100-126 mg/dl) and/or impaired glucose tolerance (IGT; blood glucose at 120 min of oral glucose tolerance test [OGTT] 140-200 mg/dl) according to the criteria of the American Diabetes Association (1). Exclusion criteria included overt diabetes mellitus or antidiabetic medication, BMI > 45 kg/m², current pregnancy or breastfeeding, serious disease (e.g. symptomatic coronary heart disease, serious symptomatic malignant disease, severe liver or kidney disease, systemic infection, severe mental illness), drug abuse, treatment with systemic corticosteroids. Claustrophobia, metal implants and excessive body circumference were deemed as contraindication of magnetic-resonance imaging, only. Participants underwent a detailed metabolic phenotyping including measurement of anthropometric parameters, body composition, liver fat content, and an OGTT.

Study subjects completed handwritten food records, noting the start and end of each eating event, the amount and kind of food consumed, for four days including three working days and one free day to reflect their dietary habits. They did not receive any dietary recommendations or restrictions before completing the food records.

### 2.2. Anthropometric measurements

For the assessment of BMI, body weight was measured using a digital scale and body height was measured using a stadiometer. Waist and hip circumferences were measured using a metric tape. Fat mass was analysed using bioelectrical impedance analyser Nutriguard MS (Data Input GmbH, Pöcking, Germany). Total and visceral adipose tissue (VAT) compartments of the trunk were assessed by magnetic resonance imaging (MRI). Liver fat content was determined by localized proton magnetic resonance spectroscopy (^1^H-MRS) applying a single-voxel stimulated echo acquisition mode sequence in the posterior hepatic segment 7 as described (27).

### 2.3. OGTT, blood sample analyses, and assessment of glycaemic indices

A standardized 2-hour OGTT was conducted after an overnight fast (>10 h since last food intake) starting between 8:00 and 9:00. After a fasting blood sampling, participants ingested standardised glucose solution (75 g glucose in 300 ml water; Accu-Chek Dextro O.G.T., Roche), and further blood samples were drawn from the forearm vein 30, 60, 90, and 120 min after glucose intake. Routine serum parameters were determined using an automated analyzer (ABX Pentra 4000; ABX, Montpellier, France). Glucose concentrations were analysed locally by certified laboratories using the glucose oxidase method. Serum insulin and C-peptide concentrations were determined by immunoassays with the ADVIA Centaur XP Immunoassay System (Siemens Healthineers, Erlangen, Germany) or with a Tecan infinite M200 microplate reader (Tecan, Maennedorf, Switzerland).

Glucose and insulin values in OGTT were used to calculate indices of insulin sensitivity (homeostasis model assessment of insulin resistance [HOMA-IR] and Matsuda index of insulin sensitivity [ISI Matsuda]), as well as insulin secretion (insulinogenic and disposition indices). HOMA-IR was calculated as (Gluc_0_ (mmol/l) x Ins_0_ (mU/l)) / 22.5. The insulinogenic index was calculated as Ins_30-0_ (mU/l) /Gluc_30-0_ (mg/dl) (30). ISI Matsuda and the disposition index, which reflects the relation between β-cell secretion capacity and insulin sensitivity, were assessed as described (31; 32).

### 2.4. Analysis of eating timing pattern

The handwritten food records were digitalized using the PRODI software version 4.5 (NutriScience GmbH, Hofstetten-Flüh, Switzerland) by an experienced nutritionist. The digital versions were used to assess time, energy intake, and food composition (carbohydrates, protein, fat, and fiber) of each eating event for each of the four consecutive days (three working days and one weekend day) for all participants. Incomplete dietary records were excluded from the analysis. The temporal eating pattern was characterised by the assessment of (i) eating timing itself (i.e., timing of the first and last eating event, eating window duration), (ii) calorie distribution across the day (i.e., time of caloric midpoint and calories in the first and last meal), and (iii) eating frequency (i.e., number of eating events per day) as described in details previously (6). In brief, an eating event was defined by the following rules: (1) at least 50 kcal, and (2) a time gap to another food or meal of at least 15 minutes (33; 34). Start and end of eating were defined as the start of the first caloric event of the day and the end of the last caloric event of the day, respectively. Eating window duration was calculated as the time difference from the start of the first eating event to the end of the last eating event, and the caloric midpoint (CM) was assessed as the time when the 50% of total calorie intake was reached (33).

To assess the timing and energy intake in the first and last meals, eating events were classified as a meal or as a snack, based on the time of the day, nutrient density, and regularity as described (6). In brief, an eating event was defined as a “meal” if it had > 15.0 % of daily energy intake (35), was consumed at the main meal times between 08:00-10:00, 12:00 – 14:00, and 18:00 – 20:00 (typical breakfast, lunch, and dinner times in Germany) (36), and was regular.

### 2.5. Statistical Analysis

Statistical analyses were carried out using SPSS 28.0 (IBM SPSS, Chicago, IL). Data were expressed as mean (SD). Associations between eating timing parameters and glycaemic and anthropometric traits were examined using multivariable linear regression models to estimate a standardized regression coefficient (β) and 95% confidence interval (CI). Primary models were adjusted for potential confounders, including age, sex, and daily energy intake. Some models were additionally adjusted for macronutrient composition and body fat. Further, for glycaemic and anthropometric traits, an unstandardized regression coefficient B was shown to estimate their alterations per each 1-unit change in original predictor scale (e.g. per hour for the eating time). Group comparison was conducted by Student’s t test or by Mann-Whitney U test. Statistical significance was defined as p < 0.05.

## 3. Results

### 3.1. Clinical characteristics of participants

Eating pattern was assessed in 297 individuals (age: 59.5 (8.9) years, BMI: 31.3 (6.1) kg/m², 106 males and 191 females) who completed food records and provided full metabolic data. Body fat content was 34.9 (8.1) %. Visceral fat accounted for 24.4 (9.2) % of total fat, and liver fat content was 9.12 (8.00) %. Average fasting glucose was 106.7 (9.5) mg/dL, and HbA1c of 5.7 (0.4) %. Anthropometric measures and parameters of glucose metabolism are shown in **Table 1**.

**Table 1.** Clinical characteristics of study participants.

| **Parameters** | **All** |
| --- | --- |
| Male / female | 106 / 191 |
| Age [years] | 59.5 (8.9) |
| **Anthropometric parameters** |  |
| BMI [kg/m<sup>2</sup>] | 31.3 (6.1) |
| Waist circumference [cm] | 101.8 (14.7) |
| Total body fat [%] | 34.9 (8.1) |
| Visceral fat [% of total fat] | 24.4 (9.1) |
| Liver fat [%] | 9.12 (8.00) |
| **Blood pressure** |  |
| Systolic blood pressure [mmHg] | 136.2 (15.1) |
| Diastolic blood pressure [mmHg] | 88.2 (10.3) |
| **Lipid parameters** |  |
| Triglycerides [mg/dl] | 139.3 (76.0) |
| Total cholesterol [mg/dl] | 211.9 (36.7) |
| HDL cholesterol [mg/dl] | 54.1 (15.2) |
| LDL cholesterol [mg/dl] | 137.6 (32.6) |
| **Glycaemic parameters** |  |
| Fasting glucose [mg/dl] | 106.7 (9.5) |
| AUC glucose [mg/dl*min] | 19839 (3212) |
| Fasting insulin [mU/l] | 14.4 (8.6) |
| AUC insulin [mU/l*min] | 13795 (9242) |
| HOMA-IR [AU] | 3.82 (2.32) |
| ISI Matsuda [AU] | 2.63 (1.65) |
| Insulinogenic index [AU] | 1.23 (1.14) |
| Disposition index [AU] | 2.67 (2.90) |
| HbA1c [%] | 5.7 (0.4) |

Data are shown as mean (SD).

### 3.2. Eating timing pattern of participants

Daily distribution of eating events is shown in **Figure S1.** Temporal eating pattern of study participants, including eating timing, calorie distribution across the day, and eating frequency, is shown in **Table 2**. Participants consumed 4.07 (0.92) meals per day, with an eating start (ES) at 08:08 (01:24) and an eating end at 20:06 (02:13), resulting in an eating window duration of 12:09 (01:49) hours (**Table 2**). Energy intake was 27.7 (10.1) % of daily calories in the first and 33.8 (11.3) % in the last meal, while the CM was estimated at 15:19 (02:04) (**Table 2**).

**Table 2.** Dietary composition and meal timing pattern of study participants.

| Parameters | All |
| --- | --- |
| **Daily dietary composition** |  |
| Daily calorie intake [kcal] | 1941 (547) |
| Carbohydrates [g] | 198.9 (65.1) |
| Carbohydrates [EN%] | 42.6 (7.5) |
| Protein [g] | 83.6 (24.7) |
| Protein [EN%] | 18.2 (4.6) |
| Fat [g] | 80.6 (30.5) |
| Fat [EN%] | 36.7 (7.6) |
| Fibre [g] | 24.0 (8.3) |
| Fibre [EN%] | 2.56 (1.02) |
| **Eating timing** |  |
| Eating window [hh:mm] | 12:09 (01:49) |
| Start of first eating event [hh:mm] | 08:09 (01:24) |
| End of last eating event [hh:mm] | 20:06 (02:13) |
| **Daily calorie distribution** |  |
| Caloric midpoint [hh:mm] | 15:19 (02:04) |
| Calories in first meal [%] | 27.7 (10.1) |
| Calories in last meal [%] | 33.8 (11.3) |
| **Eating frequency** |  |
| Number of eating events | 4.07 (0.92) |

Data are shown as mean (SD).

Eating timing parameters were tightly interrelated. In particular, ES was linked to the daily calorie distribution, showing positive associations with caloric midpoint (CM, β = 0.332, p < 0.001) and the percentage of energy in the first (β = 0.178, p = 0.002) and last meal (β = 0.167, p = 0.004). ES was negatively associated with eating duration (β = −0.582, p < 0.001) and number of meals per day (β = −0.182, p = 0.002). This indicates that individuals who started eating later had shorter eating windows and consumed fewer meals per day, but the individual meals, at least the first and last ones, tended to be larger (**Table S1**). Further, CM was positively associated with calorie intake during the last meal, total daily calorie intake, and the duration of the daily eating window, and negatively associated with calorie intake during the first meal. This suggests that individuals with a late CM tended to have a longer eating window and to consume more evening and daily calories (**Table S1**).

### 3.3. Association of eating start and glucose metabolism parameters

In an association analysis between eating timing characteristics and glycaemic traits, most associations were found for ES. In non-adjusted models, ES showed positive associations with fasting and 120-min insulin, AUC insulin, HOMA-IR, and a negative association with ISI Matsuda (**Figure 1A**). To account for potential confounders, the model was adjusted for age, sex, and daily energy intake, but these associations remained significant (**Figure 1B**). In adjusted models, each later hour of ES was associated with a 1.08 mU/l higher fasting insulin (95% CI, 0.40 to 1.77 mU/l), a 16.86 mU/l higher 120-min insulin (95% CI, 5.62 to 28.10 mU/l), a 1147 mU/l*min higher AUC insulin (95% CI, 390 to 1904 mU/l*min), a 0.32 units higher HOMA-IR (95% CI, 0.14 to 0.50), and a −0.18 units lower ISI Matsuda (95% CI, −0.32 to −0.04) (**Table 3**). This suggests that individuals with a late ES tend to have higher insulin levels and lower insulin sensitivity, independently of the energy intake. Additional adjustment for the daily intake of carbohydrates, fats, and proteins did not affect these relationships (**Table S2**).

**Figure 1.**
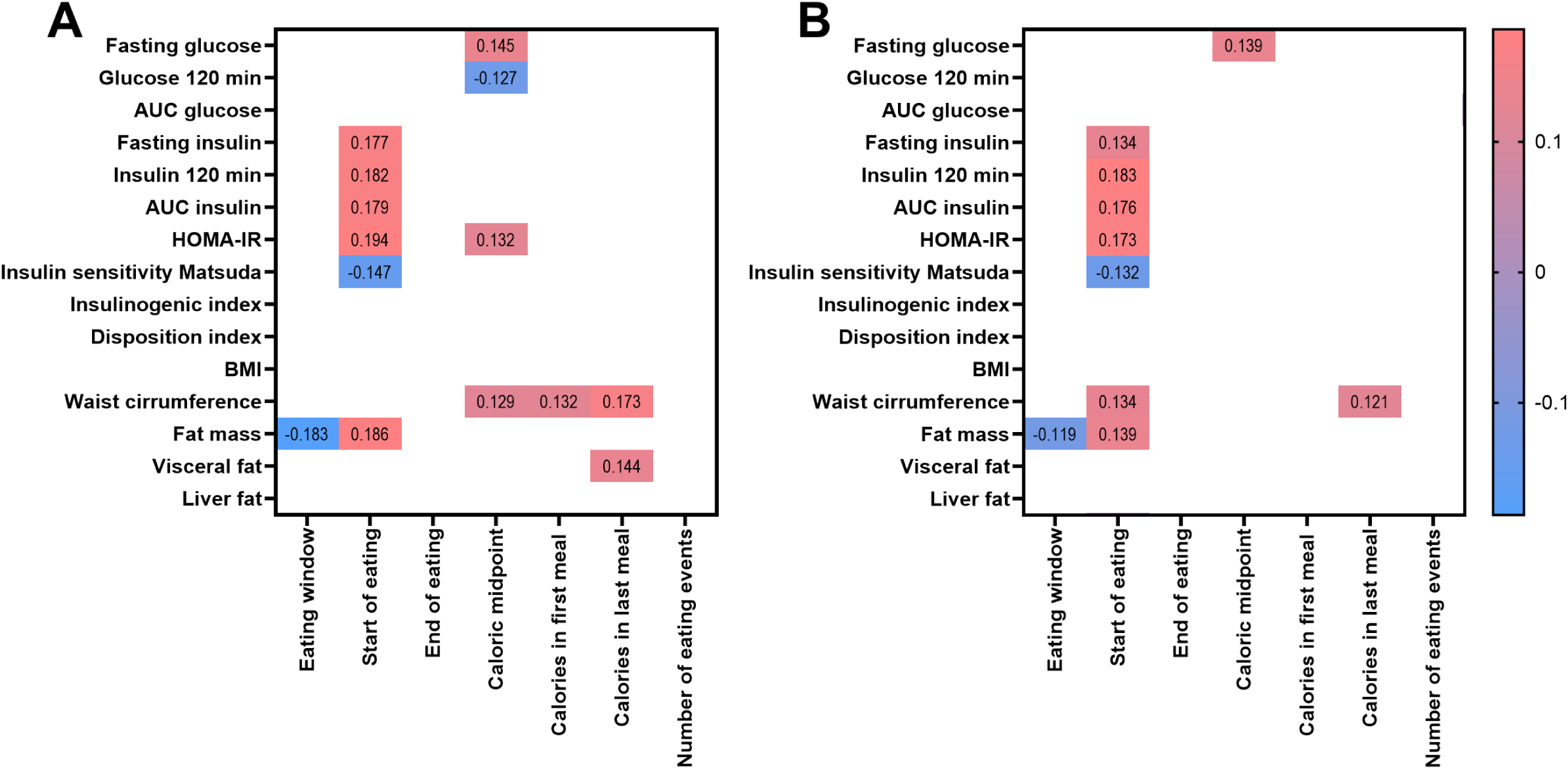
Association of eating timing, glycaemic and anthropometric parameters. Heatmap represents beta-coefficient values calculated using linear regression models without (A) and with (B) adjustment for sex, age, and daily energy intake. Eating timing parameters were averaged over four days for each person. Intensity of colour represents the strength of the association (beta-coefficients), whereas red indicates a positive and blue a negative association.

**Table 3.**
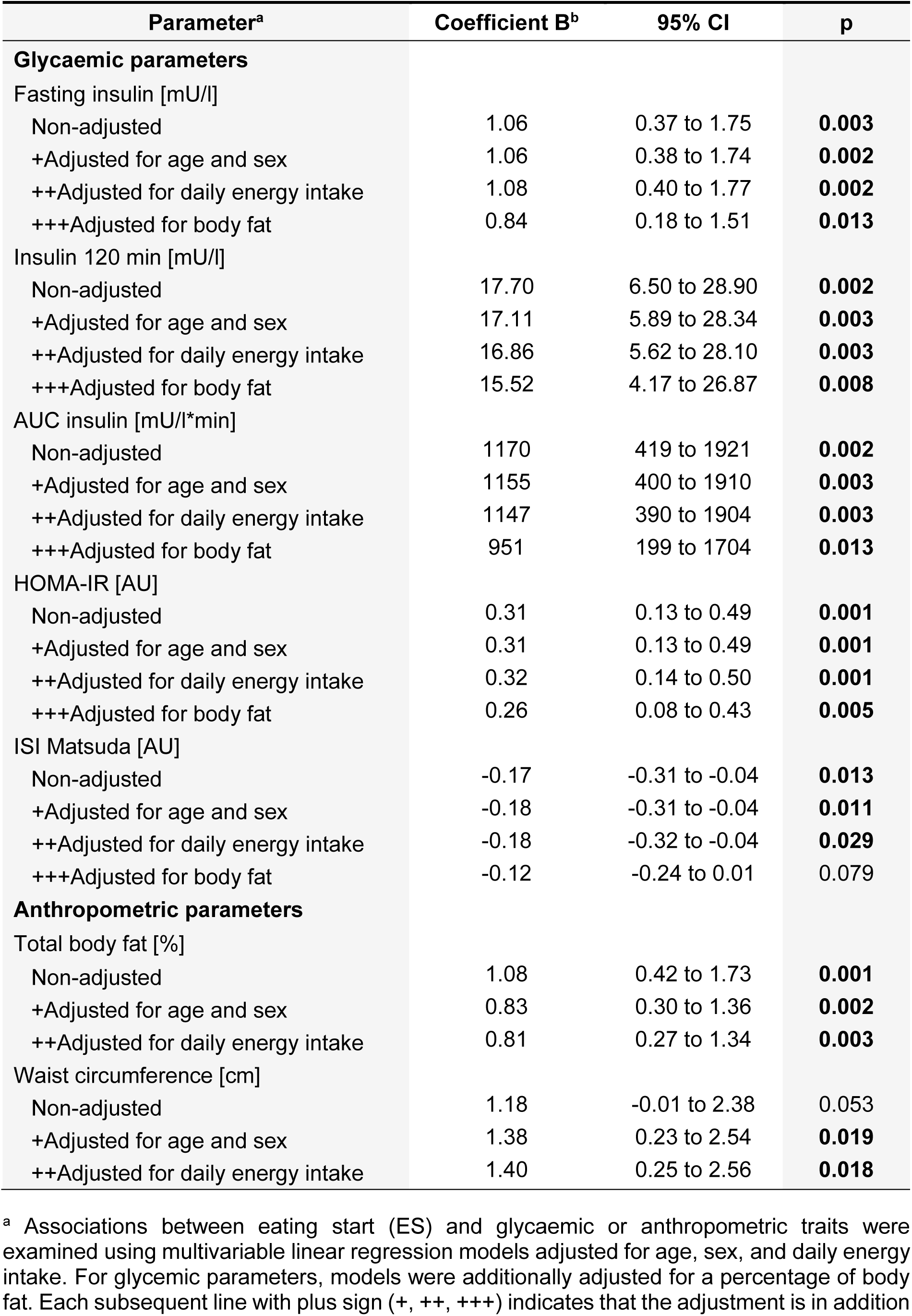
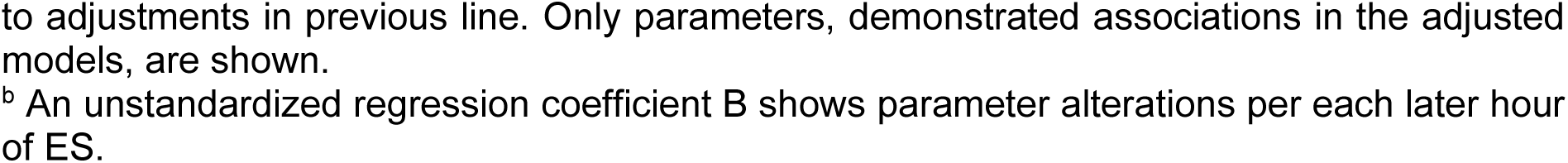
Associations of eating start with insulin levels, insulin sensitivity, and body fat with adjustments.

| Parameter<sup>a</sup> | Coefficient B<sup>b</sup> | 95% CI | p |
| --- | --- | --- | --- |
| **Glycaemic parameters** |  |  |  |
| Fasting insulin [mU/l] |  |  |  |
| Non-adjusted | 1.06 | 0.37 to 1.75 | **0.003** |
| +Adjusted for age and sex | 1.06 | 0.38 to 1.74 | **0.002** |
| ++Adjusted for daily energy intake | 1.08 | 0.40 to 1.77 | **0.002** |
| +++Adjusted for body fat | 0.84 | 0.18 to 1.51 | **0.013** |
| Insulin 120 min [mU/l] |  |  |  |
| Non-adjusted | 17.70 | 6.50 to 28.90 | **0.002** |
| +Adjusted for age and sex | 17.11 | 5.89 to 28.34 | **0.003** |
| ++Adjusted for daily energy intake | 16.86 | 5.62 to 28.10 | **0.003** |
| +++Adjusted for body fat | 15.52 | 4.17 to 26.87 | **0.008** |
| AUC insulin [mU/l*min] |  |  |  |
| Non-adjusted | 1170 | 419 to 1921 | **0.002** |
| +Adjusted for age and sex | 1155 | 400 to 1910 | **0.003** |
| ++Adjusted for daily energy intake | 1147 | 390 to 1904 | **0.003** |
| +++Adjusted for body fat | 951 | 199 to 1704 | **0.013** |
| HOMA-IR [AU] |  |  |  |
| Non-adjusted | 0.31 | 0.13 to 0.49 | **0.001** |
| +Adjusted for age and sex | 0.31 | 0.13 to 0.49 | **0.001** |
| ++Adjusted for daily energy intake | 0.32 | 0.14 to 0.50 | **0.001** |
| +++Adjusted for body fat | 0.26 | 0.08 to 0.43 | **0.005** |
| ISI Matsuda [AU] |  |  |  |
| Non-adjusted | -0.17 | -0.31 to -0.04 | **0.013** |
| +Adjusted for age and sex | -0.18 | -0.31 to -0.04 | **0.011** |
| ++Adjusted for daily energy intake | -0.18 | -0.32 to -0.04 | **0.029** |
| +++Adjusted for body fat | -0.12 | -0.24 to 0.01 | 0.079 |
| **Anthropometric parameters** |  |  |  |
| Total body fat [%] |  |  |  |
| Non-adjusted | 1.08 | 0.42 to 1.73 | **0.001** |
| +Adjusted for age and sex | 0.83 | 0.30 to 1.36 | **0.002** |
| ++Adjusted for daily energy intake | 0.81 | 0.27 to 1.34 | **0.003** |
| Waist circumference [cm] |  |  |  |
| Non-adjusted | 1.18 | -0.01 to 2.38 | 0.053 |
| +Adjusted for age and sex | 1.38 | 0.23 to 2.54 | **0.019** |
| ++Adjusted for daily energy intake | 1.40 | 0.25 to 2.56 | **0.018** |

<sup>a</sup> Associations between eating start (ES) and glycaemic or anthropometric traits were examined using multivariable linear regression models adjusted for age, sex, and daily energy intake. For glycaemic parameters, models were additionally adjusted for a percentage of body fat. Each subsequent line with plus sign (+, ++, +++) indicates that the adjustment is in addition
to adjustments in previous line. Only parameters, demonstrated associations in the adjusted models, are shown. <sup>b</sup> An unstandardized regression coefficient B shows parameter alterations per each later hour of ES.

ES was also positively associated with percentage of body fat (0.81% per ES hour; 95% CI, 0.27 to 1.34%) and waist circumference (1.4 cm per ES hour; 95% CI, 0.2 to 2.6 cm), but not with BMI or liver fat (**Figure 1B**). To prove whether ES associations with insulin levels and sensitivity are mediated by the excessive body fat, we additionally adjusted the models for this parameter, but all associations remained significant (**Table 3**).

### 3.4. Associations of other eating timing parameters with glycaemic and anthropometric traits

Non-adjusted analysis of other eating timing parameters revealed several associations with glycaemic traits, most of which disappeared in adjusted models (**Figure 1, Table S3)**. Only, CM remained positively associated with fasting glucose (0.62 mg/dl per ES hour; 95% CI, 0.10 to 1.15 mg/dl) after adjusting for age, sex, and daily energy intake. Further, calory intake in the last meal remained positively associated with waist circumference, while daily eating window was negatively associated with total body fat (**Table S3**). No associations with glycaemic or anthropometric traits were found for the time of eating end and the daily number of meals (**Figure 1**).

### 3.5. Comparison of the glycaemic and anthropometric parameters in individuals with early vs late start of eating

Based on the observed associations between ES and glycaemic parameters, all participants were categorized into groups with early ES (before 08:09, which was the average ES in the study cohort) and with late ES (after 08:09). The groups did not differ in BMI, waist circumference, visceral and liver fat, and blood lipids, but individuals with late ES showed 2.7 (0.9) % higher body fat (p=0.004) and were on average 1.9 (1.0) years older (p=0.018) (**Table 3, Table S4**).

Participants with late ES ended to eat at a similar time of day as individuals with early ES, resulting in an average 1 h 42 min shorter eating window (p<0.001) and a 42 min later CM (p=0.003) (**Table 4**). Daily food composition and calorie intake did not differ between groups, whereas percentage of energy within the first meal was slightly larger in individuals with late ES (p=0.041) (**Table S5**). In accordance with the association analysis, in the OGTT, participants with late ES showed higher insulin (p=0.029) and C-peptide (p=0.034) levels at 120 min of test, and a tendency towards higher fasting insulin (p=0.088), HOMA-IR (p=0.070), and lower ISI Matsuda (p=0.096) compared to the early ES group, whereas the glucose values were similar in both groups (**Figure 2**, **Table 4**).

**Figure 2.**
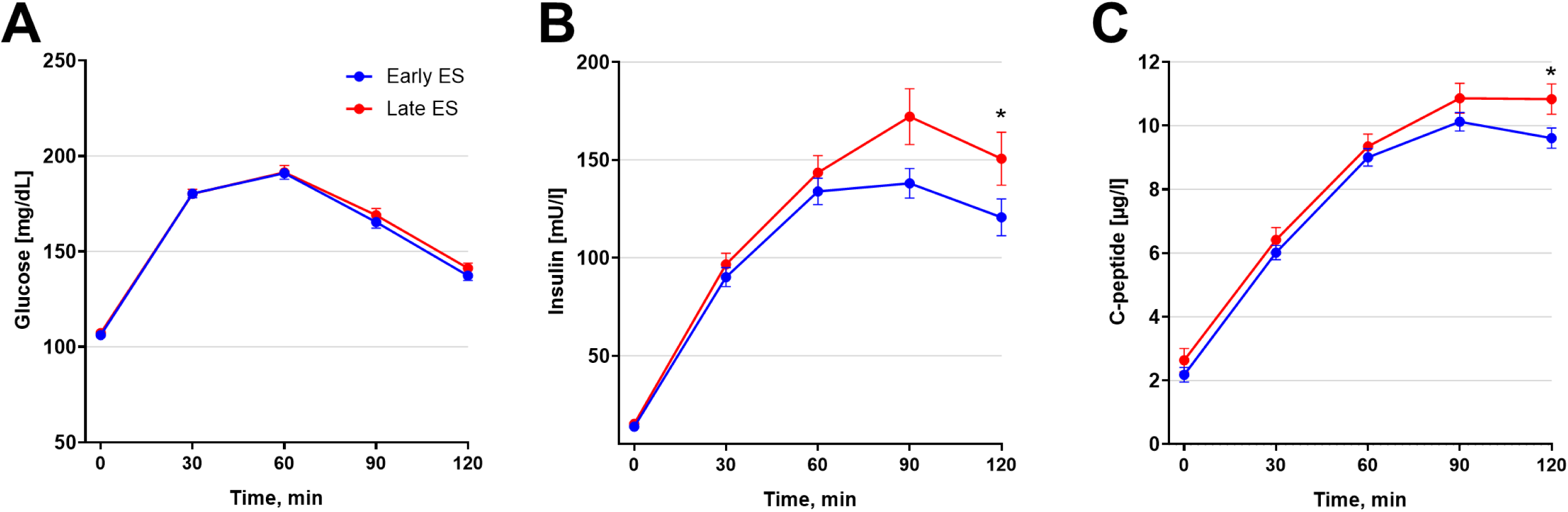
Glucose and insulin values in OGTT in individuals with early vs late start of eating. Glucose (A), insulin (B), and C-peptide (C) values in OGTT are presented in individuals with early (blue) and late (red) start of eating. Data are shown as mean ± SEM. * P<0.05 in the Student’s t test. ES, eating start.

**Table 4.** Characteristics of individuals with early vs late start of eating.

| Parameters | Early eating start | Late eating start | P |
| --- | --- | --- | --- |
| Male / female | 65 / 95 | 41 / 96 |  |
| Age [years] | 58.6 (8.6) | 60.5 (9.2) | **0.018** |
| **Anthropometric parameters** |  |  |  |
| BMI [kg/m<sup>2</sup>] | 31.3 (6.3) | 31.3 (6.0) | 0.994 |
| Waist circumference [cm] | 101.6 (14.3) | 102.1 (15.2) | 0.781 |
| Fat mass [%] | 33.6 (8.6) | 36.3 (7.3) | **0.004** |
| Visceral fat [% of total fat] | 24.4 (9.4) | 24.4 (8.9) | 0.743 |
| Liver fat [%] | 8.84 (8.03) | 9.42 (7.99) | 0.466 |
| **Glycaemic parameters** |  |  |  |
| Fasting glucose [mg/dl] | 106.2 (9.3) | 107.2 (9.6) | 0.366 |
| Glucose 120 min [mg/dl] | 137.2 (30.9) | 141.2 (30.8) | 0.270 |
| AUC glucose [mg/dl*min] | 19752 (3201) | 19942 (3233) | 0.612 |
| Fasting insulin [mU/l] | 13.8 (8.3) | 15.2 (9.0) | 0.088 |
| Insulin 120 min [mU/l*min] | 120.7 (117.2) | 150.6 (157.5) | **0.029** |
| AUC insulin [mU/l*min] | 12877 (7944) | 14851 (10469) | 0.146 |
| Fasting C-peptide [µg/l] <sup>a</sup> | 2.18 (2.60) | 2.63 (3.89) | 0.282 |
| C-peptide 120 min [µg/l] <sup>a</sup> | 9.61 (3.65) | 10.84 (4.98) | **0.034** |
| AUC C-peptide [µg/l*min] <sup>a</sup> | 931 (296) | 1001(474) | 0.372 |
| HOMA-IR | 3.62 (2.20) | 4.05 (2.44) | 0.070 |
| ISI Matsuda [AU] | 2.75 (1.64) | 2.49 (1.66) | 0.096 |
| Insulinogenic index | 1.17 (0.96) | 1.31 (1.31) | 0.587 |
| Disposition index [AU] | 2.64 (2.33) | 2.70 (3.45) | 0.460 |
| HbA1c [%] | 5.7 (0.4) | 5.7 (0.3) | 0.663 |
| **Eating timing** |  |  |  |
| Eating window [hh:mm] | 12:56 (1:34) | 11:14 (1:39) | **<0.001** |
| Start of eating [hh:mm] | 7:11 (0:43) | 9:15 (1:09) | **<0.001** |
| End of eating [hh:mm] | 20:06 (1:35) | 20:06 (2:47) | 0.411 |
| **Daily calorie distribution** |  |  |  |
| Daily calorie intake [kcal] | 1983 (586) | 1891 (496) | 0.302 |
| Caloric midpoint [hh:mm] | 14:59 (1:58) | 15:41 (2:08) | **0.003** |
| Calories in first meal [%] | 26.7 (10.4) | 28.7 (9.7) | **0.041** |
| Calories in last meal [%] | 33.1 (11.2) | 34.7 (11.4) | 0.181 |
| **Eating frequency** |  |  |  |
| Number of eating events | 4.21 (0.95) | 3.91 (0.87) | **0.006** |

Data are shown as mean (SD). Group comparison was conducted by Student's t test or by Mann-Whitney U test. <sup>a</sup> C-peptide data was available for 239 individuals.

## 4. Discussion

As the prevalence of diabetes and prediabetes continues to rise globally, understanding the relationship between eating timing and glucose metabolism becomes increasingly important, potentially offering an additional non-pharmacological approach to prevent or delay the development of T2D. In this study, we demonstrated associations of ES timing with hyperinsulinemia and insulin resistance, which represent key players in the progression of prediabetes towards T2D (37) and typically develop 10-15 years before diabetes diagnosis (38). In our study, individuals with prediabetes underwent comprehensive metabolic phenotyping, including OGTT, MRI/MRS, and documentation of their nutritional habits using food diaries. Given this comprehensive approach, the highlight of this study is the detailed characterization of individual temporal eating patterns through assessment of eating timing itself and the diurnal distribution of energy intake.

Our analysis revealed a number of associations between eating timing characteristics and glycaemic parameters, with most associations found for ES. We demonstrated that a later ES is associated with higher fasting and postprandial insulin values and lower insulin sensitivity, while no relationship was found between ES timing and glucose values. Remarkably, these associations were very robust, remaining significant after adjusting for potential cofounders - age, gender, and energy intake. While ES impact on fasting parameters (insulin, HOMA-IR) was relatively small or moderate, ES-related differences in post-challenge insulin responses in OGTT (16.86 mU/L higher 120-min insulin and 1147 mU/L*min higher AUC insulin for every hour later ES) were more pronounced. For individuals with a 3-hour later ES, this would translate to an increase of 50.58 mU/L 120-min insulin and of 3441 mU/L*min AUC insulin, representing substantial hyperinsulinemia and possibly a delayed/late insulin peaks, strongly associated with increased T2D risk (39). Considering the ES associations with HOMA-IR and ISI Matsuda, our data suggest that individuals with a late ES have more pronounced insulin resistance, which apparently results in higher compensatory hyperinsulinemia to maintain glucose control due to still sufficient beta-cell capacities. Grouping individuals based on early versus late ES supports this hypothesis, as the late ES group showed higher 120-minute insulin and C-peptide levels, despite comparable glucose values.

Importantly, in our study, later ES was also associated higher body fat, a factor, which strongly relates to obesity and might be another cofounding factor contributing to observed associations between ES and glycemic traits. Specifically, each later hour of ES was associated with a 0.81-unit higher body fat percentage after adjusting for age, sex, and energy intake. Hence, individuals with a 3-hour difference in ES timing would exhibit∼2.5% difference in percentage of body fat, which might affect metabolic and cardiovascular risks, especially if it crosses established thresholds (40). Notably, body fat content better reflects obesity and is superior to BMI for obesity-related health risks (41). Our finding on body fat aligns with published data showing an association of late eating with increased risk of obesity and higher body fat (7; 42), although we did not observe the previously reported relationship with BMI (6; 8; 15). To investigate whether ES associations with insulin levels and sensitivity are mediated by the excessive body fat, we additionally adjusted the regression models for this parameter. Remarkably, ES timing remained associated with glycemic traits, demonstrating that the effects of ES on glucose metabolism are largely independent of body fat.

Our finding aligns with previously published evidence that timing, calorie intake and composition of breakfast play an important role in glucose homeostasis and diabetes management (20; 23; 24; 43). Concerning breakfast timing, large cohort meta-analyses demonstrate an association between breakfast skipping and increased T2D risk (20; 43). T2D risk increased with every additional day of breakfast skipping, but the curve reached a plateau at 4-5 day per week, showing an increased risk of 55%, whereas this effect was only partly mediated by the weight gain (20). Cross-sectional studies revealed an association of breakfast skipping with a later chronotype, both of which contribute to poorer glycaemic control, as indicated by higher HbA1c levels (44). Several observational studies suggest that early ES at the day confers significant metabolic advantages including lower BMI, blood lipids, and better glycaemic control (3; 45). For example, data from the National Health and Nutrition Examination Survey (NHANES) demonstrated that every hour later that eating commenced was associated with approximately 0.6% higher glucose level and 3% higher HOMA-IR after adjustment for BMI and other confounders (3). A very recent study has shown that early ES is associated with lower nocturnal glucose in women with gestational diabetes (46). Notably, although in our study we did not observe difference in blood glucose, we found robust insulin changes providing clear evidence that eating timing affects glucose metabolism.

ES timing is tightly associated with calorie distribution over the day. Several studies demonstrated beneficial effects of implementing a high-calorie breakfast on glycaemic control in individuals with T2D (23; 24). In contrast, breakfast skipping is frequently accompanied by late-night dinner consumption, leading to calorie distribution towards the later part of the day. This misaligns with endogenous metabolic rhythms and can contribute to metabolic dysfunctions (47), as discussed in detail below. Interestingly, we made a similar observation in our study, showing a later CM in the late ES group, although the timing of the last eating event remained unchanged. Growing observational and experimental evidence, summarized in our and other reviews (5; 48), shows that consuming the majority of calories early in the day supports lower body weight and better glucose homeostasis compared to late or nighttime calorie intake, although detailed timing recommendations are still lacking. Notably, in our study, the calorie intake during the first meal and during the day did not differ in individuals with early and late ES. Taken together, these data suggest that the timing of ES per se, and not the calorie intake, is related to glycaemic parameters of study participants.

Interestingly, we also found that individuals with a late ES showed a shorter daily eating duration as their end of eating window did not differ from participants with an early ES. However, they showed poorer glycaemic parameters despite the shorter eating window. This finding contrasts with data from human TRE studies, reducing daily eating duration to less than 10 hours, which demonstrated beneficial effects on glucose homeostasis (49; 50). Thus, our data suggest that the timing of eating might have a stronger impact on glycaemic parameters compared to eating duration when daily calorie intake is similar in both groups. Notably, eating duration in both early and late ES groups was over eleven hours per day, which may also explain the absence of effects related to eating duration. Finally, similarly to our data, several cross-sectional or prospective human cohorts have found no link between eating window duration and body weight or metabolic traits (3; 6; 51), therefore the role of the eating duration in free-living populations remains debatable.

The next possible cofounding factor, which might affect the observed ES effects, is the daily macronutrient composition or composition of breakfast meals, which exerts profound short-term and long-term effects on glycaemic and insulinemic responses. Studies reducing breakfast carbohydrates and replacing them with proteins or fats demonstrated lower glucose, decreased glycaemic variability, improved β-cell responsiveness, and HbA1c in subjects with T2D (24; 52; 53). Based on this data, we examined in our study whether food composition might contribute to the observed ES associations with insulin levels or insulin sensitivity, but they remained significant after the additional adjustment for the macronutrient intake. We also found no differences in the macronutrient composition of the first meal or the daily average when comparing individuals with early and late ES.

Previous research has largely highlighted the CM as an eating time-related parameter, demonstrating robust associations with obesity-related traits such as BMI, waist circumference, and body fat (7; 8; 15). In addition, we recently provided evidence linking CM and insulin sensitivity in healthy subjects (6), underscoring the essential role of daily calorie distribution for glucose metabolism. In contrast to published data, our present study observed a much stronger relationship of ES with glycaemic regulation, specifically with insulin levels and sensitivity, than CM. This difference likely stems from the different study cohorts, as the CM associations were primarily observed in cohorts without focus on diabetes (7; 8; 15).

The mechanistic basis for these timing effects likely involves the circadian regulation of glucose metabolism and insulin sensitivity by endogenous clocks (54). In healthy subjects, insulin sensitivity in skeletal muscle and adipose tissue is generally higher in the early morning, facilitating rapid and efficient postprandial glucose uptake (55; 56). Similarly, pancreatic β-cells show enhanced insulin release in the morning, which effectively suppresses hepatic gluconeogenesis after breakfast (57). As the day progresses, insulin action gradually declines due to both reduced peripheral glucose uptake and diminished β-cell responses, resulting in larger glycaemic excursions in response to the same glycaemic load (57). Aligning food intake with the body’s natural metabolic rhythms – namely, early in the day – putatively leads to optimal metabolic regulation. Eating late opposes these natural rhythms and has the potential to cause circadian misalignment and metabolic dysfunction (47; 58). Remarkably, several studies showed altered circadian rhythms in individuals with T2D, with a flattened profile of insulin sensitivity and β-cell function, and elevated gluconeogenesis in the morning (59–61). Whether individuals with prediabetes also have altered metabolic rhythmicity remains insufficiently investigated (62), but if this is the case, it may help explain why our association data differ from those observed in metabolically healthy subjects. Nevertheless, our findings in early and late ES groups support the idea that individuals with prediabetes may also benefit from early eating, possibly acting as a zeitgeber and, thereby, improving their metabolic rhythmicity (5; 47).

Our study has several important strengths. The main strength is the detailed metabolic and dietary habit phenotyping in a large cohort of individuals with prediabetes, which allowed us to assess the association of eating timing with glycaemic traits – an assessment never conducted previously. Using of OGTT allowed the analysis of both fasting and postprandial metabolism, including insulin levels and the calculation of specific indices. A further strength of our study is the analysis of multiple components of the eating pattern and their potential relation to glycaemic regulation.

The prominent novelty and strength of our study is the analysis of the association of eating timing with liver fat content. Liver fat accumulation, described as MASLD, often accompanies prediabetes and T2D and strongly contributes to metabolic dysfunctions (63). Liver fat content is regulated by circadian clocks, as shown in animal models (64). Further, TRE interventions have been shown to ameliorate liver fat content in mice fed with high-fat diet (65) and in humans, with an effect size comparable to calorie restriction (66). Based on known associations of eating timing with abdominal obesity (7) and insulin sensitivity (6), we expected to observe an association of eating timing with liver fat in prediabetes. Surprisingly, no eating timing parameters showed a relation to liver fat content in our cohort. This data expands our knowledge in this field and requires further investigation in larger cohorts and diverse categories of individuals with and without metabolic dysfunctions.

Some limitations have to be also discussed when interpreting the results of this work. First, food intake and timing data were self-reported using the food diaries. Although this method is recognized as a gold standard for nutritional data collection, it does not exclude measurement errors, such as over- and underreporting. Future studies should include objective monitoring of eating behaviour using wearables or smartphone apps. Secondly, no data on sleep timing or quality, which are strongly associated with metabolism and can worsen glycaemic parameters upon the sleep deficit (67; 68), were collected. Moreover, collecting sleep time data or questionnaires to assess individual chronotypes would help to decipher how the relation between eating time and chronotype is linked to the diabetes development. Thirdly, the analysis did not consider the socioeconomic status of participants. Breakfast skipping associated with poor socioeconomic status could also explain obesity and the impaired metabolic state. Further, practicing of intermittent fasting in form of breakfast skipping could also be associated with poor metabolic state in the sense of reverse causality: as an attempt to overcome prediabetes instead of being the reason for its development. Finally, the selection bias by cutting of patients with stronger glycemic impairment (overt diabetes) has to be mentioned as a limitation.

In conclusion, our study showed that a late ES is with estimates of hyperinsulinemia and lower insulin sensitivity in prediabetes, independent of energy intake, food composition, and body fat. Our findings suggest that an earlier ES might relate to improved glucose metabolism and lower the risk of progression from prediabetes to diabetes. This non-pharmacological approach needs to be investigated in interventional studies.

## Acknowledgements

We thank all study participants for their cooperation. We gratefully acknowledge the excellent assistance of dietitians, study nurses, and lifestyle advisors involved in this study. We also thank Elwira Gliwska for her contribution to the algorithm development for the analysis of meal timing.

## Data availability

The data that support the findings of this study are available from the corresponding author upon reasonable request

## Authors’ relationships and activities

S.K. received grants from the German Diabetes Association, the Wilhelm Doerenkamp Foundation, the Almond Board of California and the California Walnut Commission, and lecture fees from Sanofi, Boehringer Ingelheim, Berlin Chemie, Lilly Deutschland and Juzo Akademie. The remaining authors declare that there are no relationships or activities that might bias, or be perceived to bias, their work.

## Contribution statement

All named authors meet the ICMJE criteria for authorship for this article, take responsibility for the integrity of the work as a whole, have reviewed the article critically for intellectual content, and have given their final approval for this version to be published. OPR is the guarantor of this work, and, as such, has full access to all the data in the study and take responsibility for the integrity of the data and the accuracy of the data analysis. All authors contributed to the methodology, the analyses, and the manuscript writing.

## Abbreviations

AUC: Area under the curve
BMI: Body mass index
CM: Caloric midpoint
ELISA: Enzyme linked immunosorbent assay
ES: Eating start
FM: Start of the first meal
HbA1c: Haemoglobin A1c
HDL: High-density lipoprotein
HIC: Hepatic insulin clearance
HOMA-IR: Homeostatic Model Assessment for Insulin Resistance
IFG: Impaired fasting glucose
IGT: Impaired glucose tolerance
ISI: Insulin sensitivity index
LDL: Low-density lipoprotein
MASLD: Metabolic dysfunction-associated steatotic liver disease
MRI: Magnetic resonance imaging
MRS: Magnetic resonance spectroscopy
OGTT: Oral glucose tolerance test
SD: Standard deviation
T2D: Type 2 diabetes
TRE: Time-restricted eating

## Supplementary material

### Supplementary Figures

**Figure S1.**
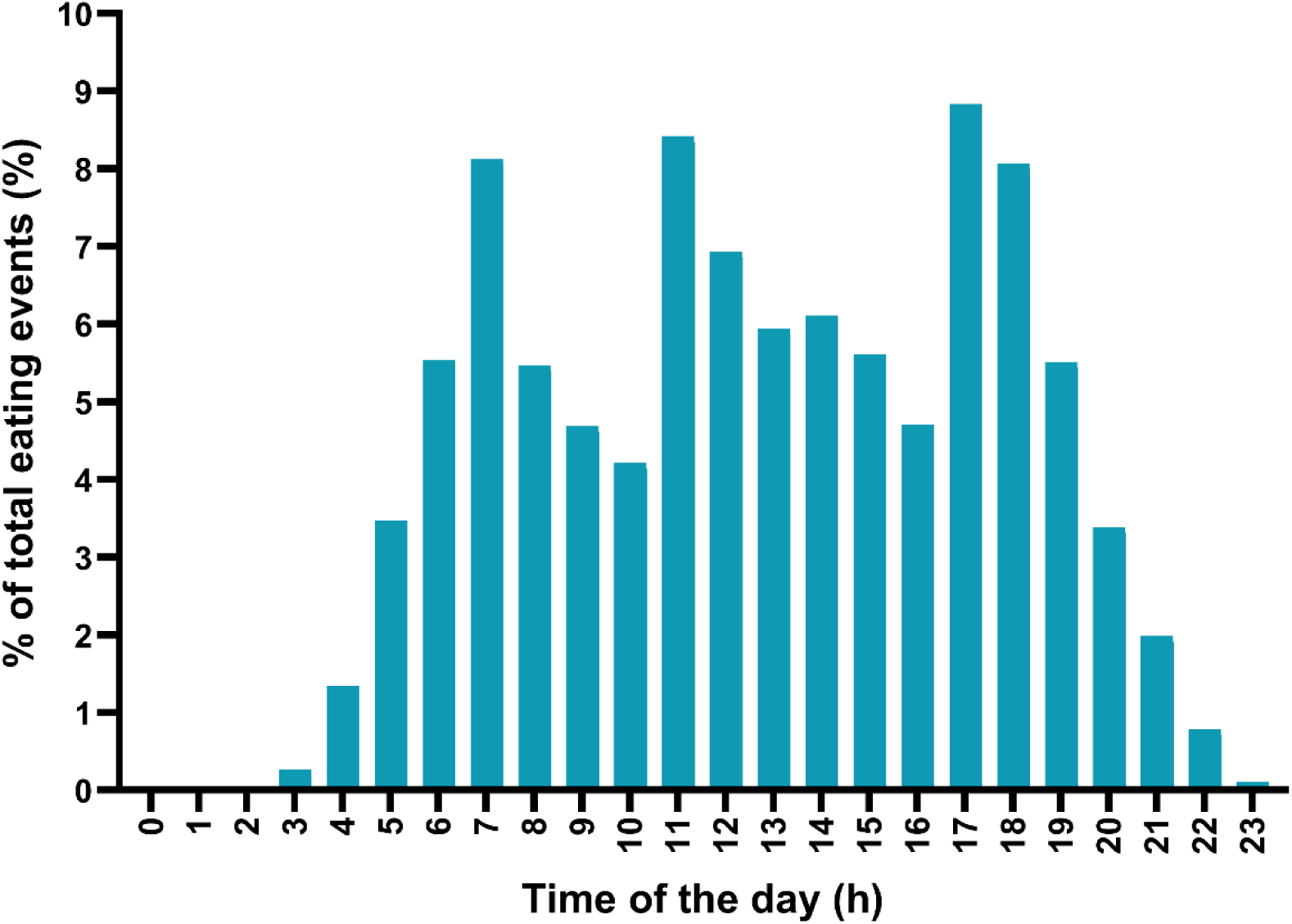
Daily distribution of eating events. Distribution of eating events is shown as percentage of all eating events in all study participants (n=297). Each number at the time axis represents an hour starting from 0 (24:00 – 1:00) and ending with 23 (23:00 – 24:00).

### Supplementary Tables

**Table S1.**
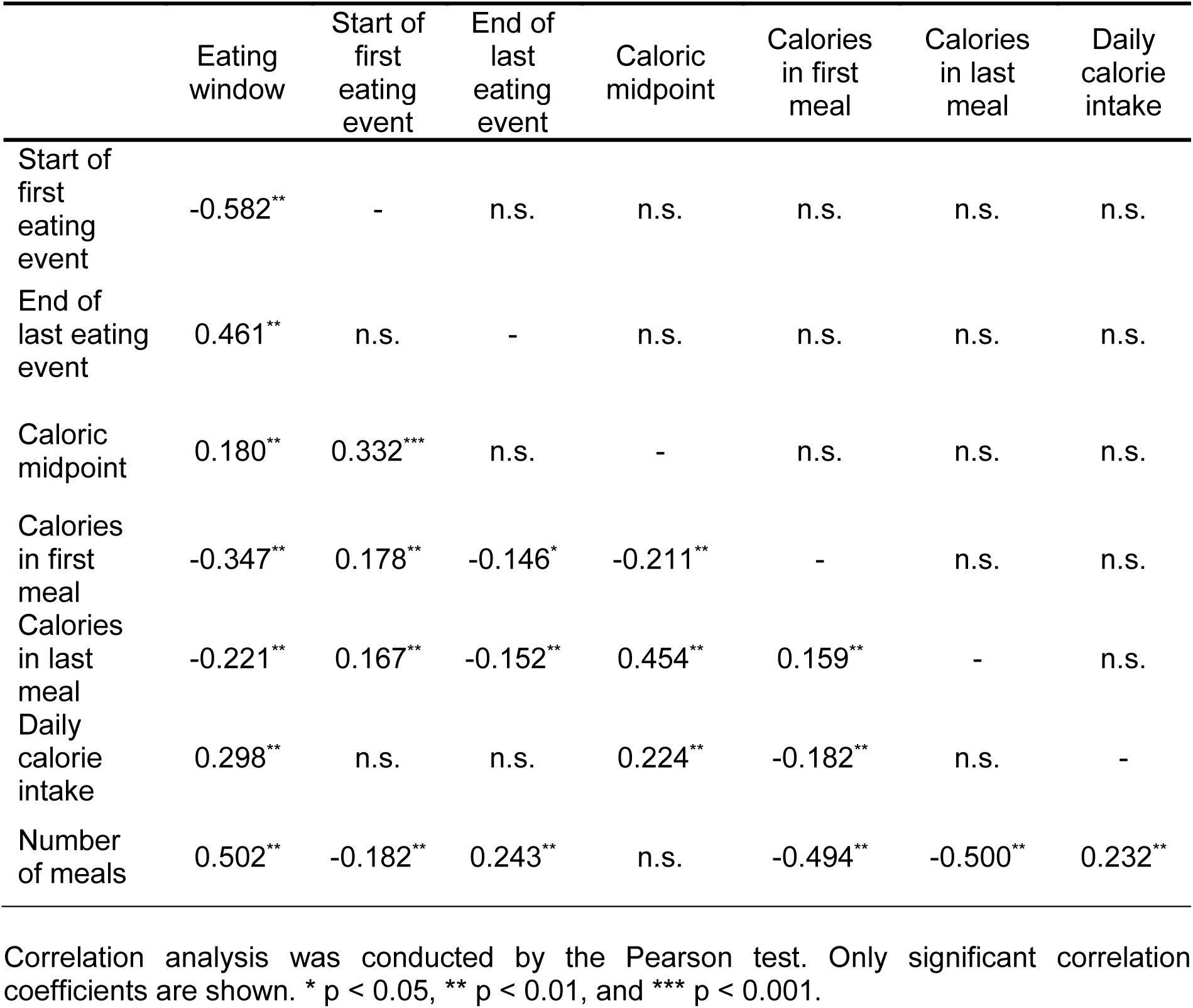
Correlation analysis of eating timing parameters and daily calorie distribution.

**Table S2.**
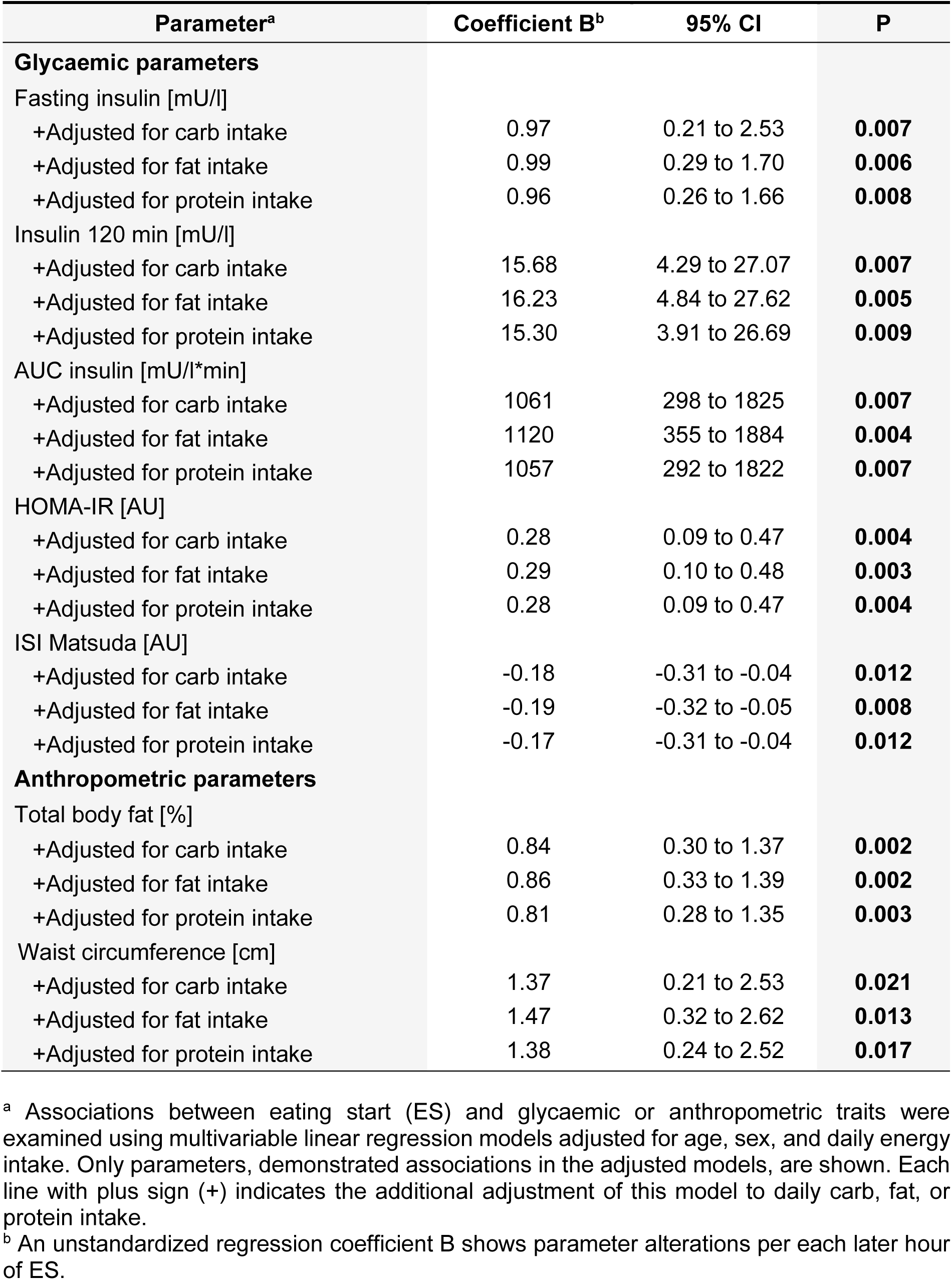
Associations of eating start with insulin levels and body fat with adjustments for macronutrient intake.

**Table S3.**
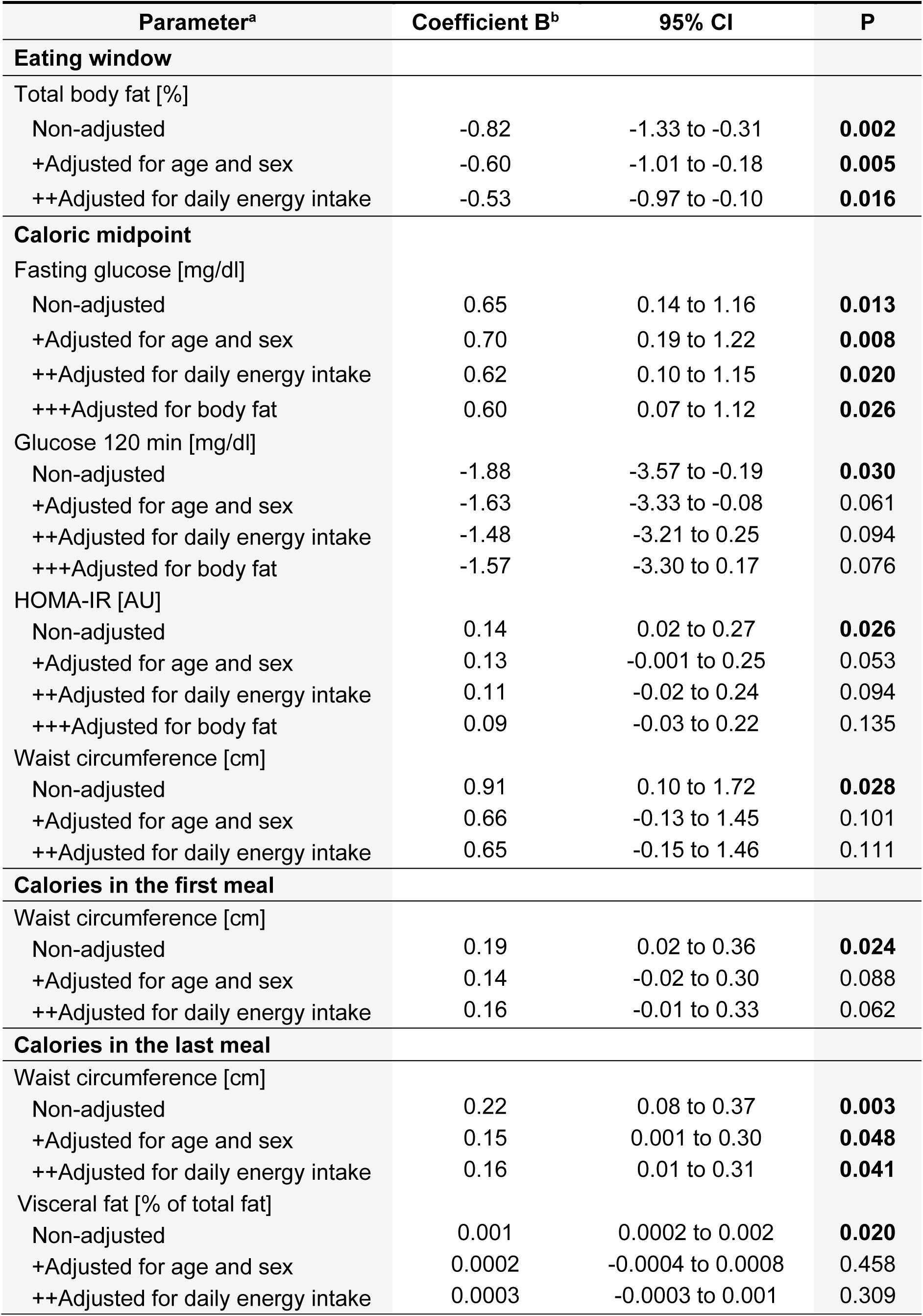
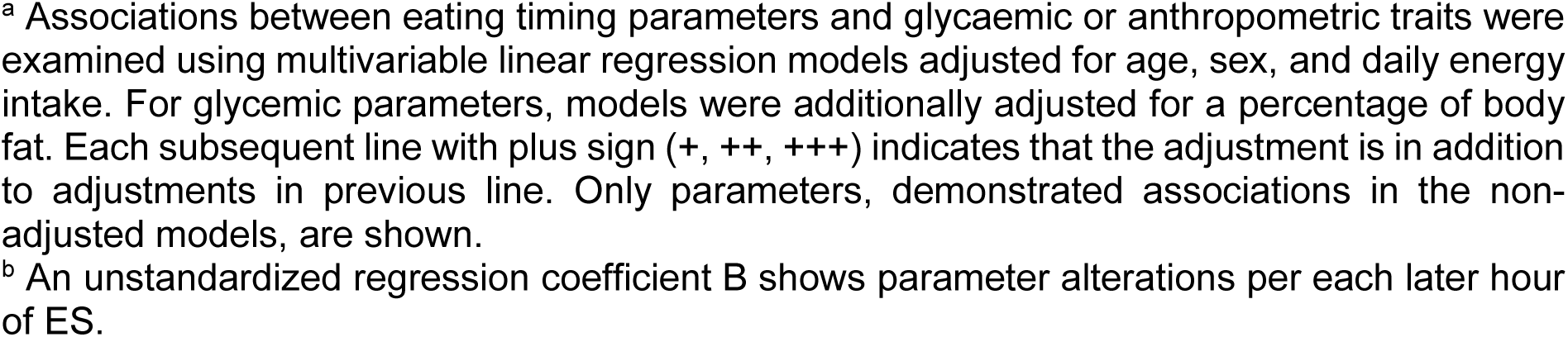
Associations of other eating timing parameters with glycaemic and anthropometric traits.

| Parameter<sup>a</sup> | Coefficient B<sup>b</sup> | 95% CI | P |
| --- | --- | --- | --- |
| **Eating window** |  |  |  |
| Total body fat [%] |  |  |  |
| Non-adjusted | -0.82 | -1.33 to -0.31 | **0.002** |
| +Adjusted for age and sex | -0.60 | -1.01 to -0.18 | **0.005** |
| ++Adjusted for daily energy intake | -0.53 | -0.97 to -0.10 | **0.016** |
| **Caloric midpoint** |  |  |  |
| Fasting glucose [mg/dl] |  |  |  |
| Non-adjusted | 0.65 | 0.14 to 1.16 | **0.013** |
| +Adjusted for age and sex | 0.70 | 0.19 to 1.22 | **0.008** |
| ++Adjusted for daily energy intake | 0.62 | 0.10 to 1.15 | **0.020** |
| +++Adjusted for body fat | 0.60 | 0.07 to 1.12 | **0.026** |
| Glucose 120 min [mg/dl] |  |  |  |
| Non-adjusted | -1.88 | -3.57 to -0.19 | **0.030** |
| +Adjusted for age and sex | -1.63 | -3.33 to -0.08 | 0.061 |
| ++Adjusted for daily energy intake | -1.48 | -3.21 to 0.25 | 0.094 |
| +++Adjusted for body fat | -1.57 | -3.30 to 0.17 | 0.076 |
| HOMA-IR [AU] |  |  |  |
| Non-adjusted | 0.14 | 0.02 to 0.27 | **0.026** |
| +Adjusted for age and sex | 0.13 | -0.001 to 0.25 | 0.053 |
| ++Adjusted for daily energy intake | 0.11 | -0.02 to 0.24 | 0.094 |
| +++Adjusted for body fat | 0.09 | -0.03 to 0.22 | 0.135 |
| Waist circumference [cm] |  |  |  |
| Non-adjusted | 0.91 | 0.10 to 1.72 | **0.028** |
| +Adjusted for age and sex | 0.66 | -0.13 to 1.45 | 0.101 |
| ++Adjusted for daily energy intake | 0.65 | -0.15 to 1.46 | 0.111 |
| **Calories in the first meal** |  |  |  |
| Waist circumference [cm] |  |  |  |
| Non-adjusted | 0.19 | 0.02 to 0.36 | **0.024** |
| +Adjusted for age and sex | 0.14 | -0.02 to 0.30 | 0.088 |
| ++Adjusted for daily energy intake | 0.16 | -0.01 to 0.33 | 0.062 |
| **Calories in the last meal** |  |  |  |
| Waist circumference [cm] |  |  |  |
| Non-adjusted | 0.22 | 0.08 to 0.37 | **0.003** |
| +Adjusted for age and sex | 0.15 | 0.001 to 0.30 | **0.048** |
| ++Adjusted for daily energy intake | 0.16 | 0.01 to 0.31 | **0.041** |
| Visceral fat [% of total fat] |  |  |  |
| Non-adjusted | 0.001 | 0.0002 to 0.002 | **0.020** |
| +Adjusted for age and sex | 0.0002 | -0.0004 to 0.0008 | 0.458 |
| ++Adjusted for daily energy intake | 0.0003 | -0.0003 to 0.001 | 0.309 |

<sup>a</sup> Associations between eating timing parameters and glycaemic or anthropometric traits were examined using multivariable linear regression models adjusted for age, sex, and daily energy intake. For glycaemic parameters, models were additionally adjusted for a percentage of body fat. Each subsequent line with plus sign (+, ++, +++) indicates that the adjustment is in addition to adjustments in previous line. Only parameters, demonstrated associations in the non-adjusted models, are shown. <sup>b</sup> An unstandardized regression coefficient B shows parameter alterations per each later hour of ES.

**Table S4.** Lipid parameters in individuals with early vs. late start of eating.

| Parameters | Early eating start | Late eating start | P |
| --- | --- | --- | --- |
| Triglycerides [mg/dl] | 137.8 (70.9) | 140.9 (81.8) | 0.812 |
| Total cholesterol [mg/dl] | 213.0 (38.8) | 210.5 (34.1) | 0.553 |
| HDL cholesterol [mg/dl] | 53.5 (15.0) | 54.8 (15.6) | 0.467 |
| LDL cholesterol [mg/dl] | 138.8 (34.9) | 136.2 (29.9) | 0.511 |

Data are shown as mean (SD). N=297.

**Table S5.** Food composition and calorie intake in individuals with early vs. late start of eating.

| Parameters | Early eating start | Late eating start | P* |
| --- | --- | --- | --- |
| **Daily dietary composition** |  |  |  |
| Calorie intake [kcal] | 1983 (586) | 1891 (496) | 0.302 |
| Carbohydrates [g] | 205.3 (66.3) | 191.4 (63.0) | 0.062 |
| Carbohydrates [EN%] | 43.0 (7.5) | 42.2 (7.6) | 0.375 |
| Protein [g] | 85.7 (28.0) | 81.2 (20.0) | 0.353 |
| Protein [EN%] | 18.2 (5.0) | 18.1 (4.0) | 0.954 |
| Fat [g] | 82.0 (32.4) | 78.9 (28.2) | 0.748 |
| Fat [EN%] | 36.2 (7.3) | 37.2 (7.8) | 0.271 |
| Fibre [g] | 25.0 (8.4) | 22.7 (8.1) | **0.024** |
| Fibre [EN%] | 2.60 (0.87) | 2.52 (1.17) | 0.300 |
| **Dietary composition of the first meal** |  |  |  |
| Calorie intake [kcal] | 488 (202) | 509 (218) | 0.524 |
| Calorie intake [% of daily intake] | 26.7 (10.4) | 28.7 (9.7) | **0.041** |
| Carbohydrates [g] | 57.5 (26.1) | 56.9 (27.0) | 0.902 |
| Carbohydrates [EN%] | 48.8 (12.0) | 46.9 (11.6) | 0.171 |
| Protein [g] | 20.2 (11.8) | 20.8 (10.0) | 0.416 |
| Protein [EN%] | 16.4 (5.6) | 16.6 (4.9) | 0.981 |
| Fat [g] | 18.8 (11.4) | 20.7 (12.4) | 0.210 |
| Fat [EN%] | 31.8 (11.2) | 33.6 (12.3) | 0.176 |
| Fibres [g] | 7.29 (3.88) | 7.07 (3.95) | 0.527 |
| Fibres [EN%] | 3.00 (1.26) | 2.82 (1.25) | 0.155 |

Data are shown as mean (SD).

